# Wearable Devices Detect Physiological Changes that Precede and Are Associated with Symptomatic and Inflammatory Rheumatoid Arthritis Flares

**DOI:** 10.1101/2025.09.24.25336585

**Authors:** Pragya Sharma, Matteo Danieletto, Jessica K Whang, Kyle Landell, Drew Helmus, Bruce E. Sands, Mayte Suarez-Farinas, Percio S. Gulko, Robert P. Hirten

## Abstract

Physiological parameters are altered in rheumatoid arthritis (RA). We evaluated whether changes in physiological metrics, collected from wearable devices, identify and precede the development of both symptomatic and inflammatory RA flares. Participants with RA answered daily disease activity surveys and provided laboratory assessments of inflammatory activity. They wore an Apple Watch (n=35), Fitbit (n=17), or Oura Ring (n=3) collecting heart rate (HR), resting heart rate (RHR), heart rate variability (HRV), and steps. Linear mixed effect models were used to associate HR, RHR and steps with flare and remission periods. Cosinor mixed effect models assessed circadian features of HRV. Mixed effect logistic regression models evaluated changes in physiological metrics prior to the onset of flares. The study enrolled 53 participants (88.7% female) with a mean age of 51.1 (SD 15.2) years. Each contributed a mean of 105 (SD 97) days of data. Mean steps were lower, while mean nighttime HR was higher during symptomatic periods, compared to periods of symptomatic remission. Means daily HR, daytime HR, nighttime HR, and RHR were higher during periods of inflammatory flares, compared to inflammatory remission. Circadian features of HRV differentiated inflammatory and symptomatic flares from remission. All metrics were altered up to 4 weeks prior to inflammatory and symptomatic flare development. This suggests the potential use of wearable devices for disease monitoring and flare prediction.

## Introduction

Rheumatoid arthritis (RA) is a chronic immune mediated inflammatory disease characterized by inflammation of the joints and symptoms such as joint pain, swelling, stiffness and reduced mobility.^1^ Its management is challenging due to the unpredictable nature of disease flares. Furthermore, there can be discordance between inflammation and symptoms, and between physician assessed and patient reported RA activity.^2^ Monitoring modalities are currently limited to single time point infrequent clinical visits, laboratory testing and clinician-dependent or patient reported assessments.^3,4^

Wearable devices are commonly used and accepted by the general population and offer a novel means to assess physiological parameters passively, frequently and outside of the clinical setting. Through remote patient monitoring, they are frequently employed for the monitoring of medical conditions such as diabetes and heart failure, where they have demonstrated improved disease outcomes.^5,6^ There is increasing interest in their use to monitor inflammatory diseases.^7^ However, in RA there are limited studies utilizing wearable devices to monitor disease activity, with most of them focusing on changes in physical activity. One study of 155 participants with either RA or axial spondyloarthritis demonstrated that patient reported flares were associated with less physical activity as measured by activity trackers, and that changes in physical activity patterns associated with patient reported flares.^8^ A study of 85 participants with RA who wore a wearable activity tracker for 7 days found lower total daily exercise and steps in those with moderate to high symptomatic disease activity.^9^ Furthermore, it has been shown that digital health measure can augment patient reported outcomes. In a study of 30 people with moderate-severe RA compared to 30 healthy controls a machine learning framework incorporating symptoms derived from an iPhone guided test, actigraphy and heart sensor data was shown to distinguish RA status and severity, as well as those with RA from healthy participants.^10^

However, chronic inflammatory diseases like RA have altered autonomic nervous system parameters. Heart rate variability (HRV) is a measure of small-time differences between each heartbeat and is an indirect measure of autonomic nervous system function.^11^ In general, lower HRV reflects increased sympathetic nervous system activity. Individuals with active RA have been shown to have lower HRV compared to those with inactive disease.^12^ Similarly, higher heart rate (HR) and resting heart rates (RHR) have been observed in RA compared to healthy controls, reflecting this autonomic dysfunction.^13,14^ However, these studies were conducted based on single timepoint or limited assessments and not via wearable devices or collected longitudinally. Many wearable devices measure such metrics in a near continuous manner, presenting an opportunity to monitor physiological parameters that relate to underlying RA activity, and thus have the potential to track and predict disease activity.

The aim of our study is to assess how longitudinally collected physiological metrics from wearable devices can identify both inflammatory and symptomatic RA activity. Furthermore, we evaluated whether these metrics change prior to the onset of inflammatory and symptomatic RA flares, exploring their future potential to predict subsequent flare events.

## Results

A total of 53 participants enrolled in the study and contributed wearable and either laboratory or survey data across 6 states in the United States (**Table 1**) with 50 participants having at least one day of either symptom or inflammatory data overlapping with their wearable metrics. The mean age of participants was 51.09 (SD 15.22) years old, with 47 (88.7%) participants being female. The mean duration of disease was 9.50 (SD 7.02) years.

**Table 1.** Demographic information for study participants.

|  | Overall Cohort (N=53) |
| --- | --- |
| Age, mean (SD) | 51.1 (15.2) |
| Sex, female (%) | 47 (88.7) |
| BMI, mean (SD) | 29.0 (7.8) |
| Race |  |
| Asian | 2 (3.8%) |
| Black | 12 (22.6%) |
| White | 31 (58.5%) |
| Missing Variable | 8 (15.1%) |
| Ethnicity |  |
| Hispanic | 15 (28.3%) |
| Not Hispanic | 36 (67.9%) |
| Missing Variable | 2 (3.8%) |
| Smoking Status |  |
| Current | 1 (1.9%) |
| Never | 36 (67.9%) |
| Past Smoker | 16 (30.2%) |
| Reported Medical Conditions |  |
| Asthma | 11 (20.8%) |
| Hypertension | 6 (11.3%) |
| Fibromyalgia | 2 (3.8%) |
| Cancer | 2 (3.8%) |
| Diabetes | 2 (3.8%) |
| Thyroid Disease | 2 (3.8%) |
| Autoimmune hepatitis | 1 (1.9%) |
| Lupus | 1 (1.9%) |
| Sjogren's Disease | 1 (1.9%) |
| Multiple Sclerosis | 1 (1.9%) |
| Sarcoidosis | 1 (1.9%) |
| RA Medications Use at Enrollment |  |
| Corticosteroids | 5 (9.4%) |
| Biologic Agents | 20 (37.7%) |
| Small Molecules | 4 (7.5%) |
| Immunomodulators | 41 (77.4%) |
| NSAIDs | 10 (18.9%) |

Thirty-five participants used an Apple Watch, 17 used a Fitbit and 3 used an Oura Ring, with one participant using both a Fitbit and Apple Watch, and one participant with both an Apple Watch and Oura Ring. The participants contributed wearable data, a mean of 154 (SD 93) days and used their wearable device an average of 16.4 (SD 4.6) hours per day. Across all 53 participants 8,183 days (142,985 hours) of wearable data is available. **Supplementary Table 1** shows data characteristics related to study duration, device compliance, survey and CRP laboratory records, flare prevalence and the missingness of data. Visualization of longitudinal wearable, survey and laboratory data for a study participant is presented in **Figure 1**.

**Figure 1.**
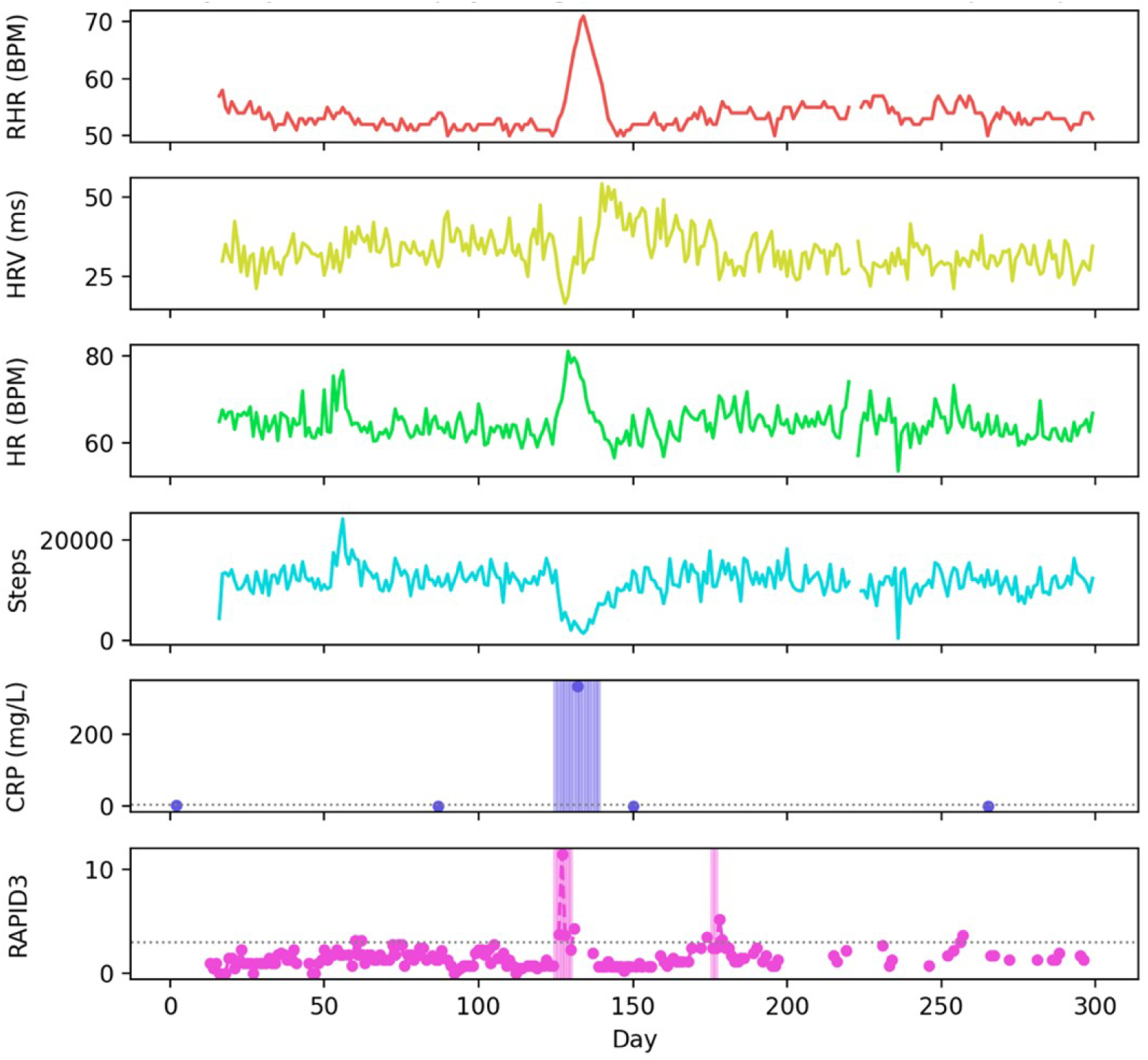
An exemplary participant’s daily trajectories are shown over the entire study duration including physiological metrics (RHR, HR, HRV RMSSD and Steps), CRP lab values and RAPID-3 daily survey scores. Detected inflammatory and symptomatic flares are shown with shaded bands in the bottom two panels. Note, an isolated RAPID-3 survey >3 is not taken as a flare, rather, at least 3 positive out of 4 surveys in a 7-day window is classified as a symptomatic flare. CRP lab values are augmented to ±7 days for inflammatory flare detection.

Wearable device data overlapping with inflammatory flare assessments were available in 38 individuals and with symptomatic flare assessments in 47 individuals. Fourteen out of 38 individuals reported elevated CRP levels (inflammatory flares) and 36 out of 47 individuals had symptomatic flares. Participants had a mean of 105 (SD 97) days with overlapping wearable and flare (symptomatic or inflammatory) data.

### Identification of Inflammatory Flares

An overview of inflammatory flare trajectories is visualized in **Supplementary Figure 1**. There were a total of 270 days of inflammatory flares and 754 days of inflammatory remission over the follow up period. Physiological metrics collected from wearable devices differentiated inflammatory periods from non-inflammatory periods. The mean daily HR, mean daytime HR, and mean nighttime HR were higher during periods of inflammatory flare (mean HR 81.42±3.03, day HR 84.26±3.10, night HR 78.63±3.43) compared to periods of inflammatory remission (mean HR 75.44±2.86, p<0.001; day HR 77.81±2.93, p<0.001; night HR 73.16±3.26, p<0.004). Similarly, RHR was higher during periods of inflammatory flare (63.21±2.22) compared to periods of inflammatory remission (58.24±2.15; p<0.0001). There was no difference in mean daily steps during periods of inflammatory flare (7,737±1,296 steps) compared to inflammatory remission (8,730±1,265 steps, p=0.09) (**Figure 2a**). **Supplementary Table 2** shows detailed flare-remission contrasts, standard error (SE) and p-values for each physiological metric and flare type.

**Figure 2.**
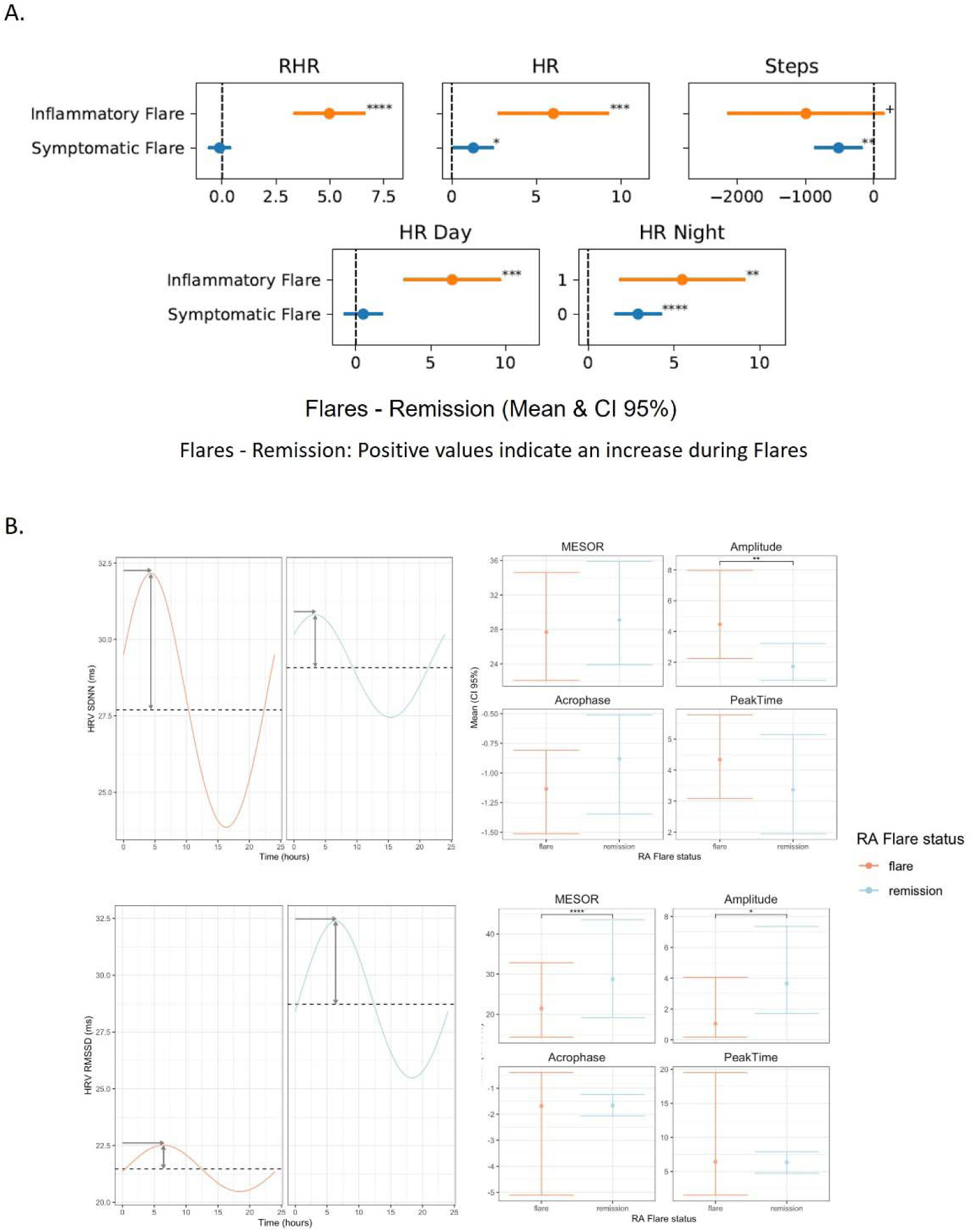
The relationship between physiological metrics measured by wearable devices and inflammatory flares. (A) The marginal means and 95% confidence intervals for resting heart rate, daily heart rate, daytime heart rate, nighttime heart rate, and steps comparing periods of inflammatory flare to inflammatory remission as well as symptomatic flare to symptomatic remission. The x-axis represents the difference between each flare and remission state, in either beats per minute or steps. (B) Circadian parameters of heart rate variability comparing periods of inflammatory flare with inflammatory remission. Stars indicate significant differences, with (+: p < 0.1; *: p < 0.05; **: p < 0.01; ***: p < 0.001, ****: p<0.0001).

There were significant differences in HRV between periods of inflammatory flare compared to periods of inflammatory remission. The circadian pattern of HRV can be described using the following parameters (1) MESOR: a rhythm-adjusted mean of the circadian pattern; (2) Amplitude: difference between the peak and the mean value giving a measure of half the extent of variation within a day; and (3) Acrophase and peak time: measures of the time when peak values occur each day. The mesor of the circadian pattern of the HRV metric root mean square of successive differences between normal heartbeats (RMSSD) was significantly lower during periods of inflammation (21.47ms 95% CI 13.28-32.74) compared to periods of inflammatory remission (28.72ms 95% CI 18.17-43.83; p<0.001). The amplitude of the circadian pattern of RMSSD is significantly different during periods of inflammatory activity (1.05ms 95% CI 0.19-4.12) compared to periods of inflammatory remission (3.66ms 95% CI 1.70-7.54, p=0.04). There was no significant difference in acrophase and peak time between the two states (p=0.98).

Similarly, there were differences in the HRV metric the standard deviation of the interbeat intervals of normal sinus beats (SDNN) based on the presence or absence of underlying inflammation. There were significant differences in the amplitude of the circadian pattern of SDNN between periods of inflammation (4.46ms 95% CI 2.25-7.98) and inflammatory remission (1.73 95% CI 0.82-3.21, p=0.006), but not in the mesor (p=0.39) and acrophase or peak time (p=0.35) (**Figure 2b**). Cosinor mixed-effect model flare-remission contrasts for mesor, amplitude, acrophase and peak time, along with p-values are shown in **Supplementary Tables 3 and 4** for both RMSSD and SDNN HRV metrics, respectively.

### Identification of Symptom Flares

An overview of the symptomatic flare trajectory is visualized in **Supplementary Figure 2**. There were a total of 1,985 days of symptomatic flares and 3,353 days of symptomatic remission in the study. The physiologic metrics collected from wearable devices differentiated periods of symptomatic flares compared to periods of symptomatic remission. The mean daily HR, and mean nighttime HR were higher during periods of symptomatic flares (mean HR 81.03±2.30, night HR 76.91±2.33) compared to periods of symptomatic remission (mean HR 79.77±2.28, p=0.04; night HR 74.01±2.30, p<0.0001). There was no significant difference in daytime HR between symptomatic flares and symptomatic remission (day HR 81.22±2.32 vs 80.72±2.31, p=0.44). There was no significant difference in RHR between the two symptom states (61.50±1.88 vs 61.62±1.88, p=0.66). During periods of symptomatic activity participants walked significantly less steps compared to periods of symptomatic remission (7,282±954 steps vs 7,797±954 steps, p=0.004) (**Figure 2a**) (**Supplementary Table 2**).

There was a significant difference in HRV between symptomatic periods compared to periods of symptomatic remission. There was a significant difference in the circadian pattern of RMSSD during symptomatic flares (Mesor 28.82ms 95% CI 21.36-38.60; acrophase -1.68 95% CI -1.93- -1.42; peak time 6.41hr 95% CI 5.41-7.37) compared to periods of symptomatic remission (Mesor 31.06ms 95% CI 23.12-41.38, p<0.001; acrophase -2.16 95% CI -2.28- -2.03, p=0.002; peak time 8.25hr 95%CI 7.77-8.71, p=0.002). Similarly, there were differences in the circadian pattern of SDNN based on symptomatic state. The amplitude, acrophase and peak time were significantly different during periods of symptomatic flares (2.47ms 95% CI 1.72-3.44; -1.85 95% CI - 2.02- -1.67; 7.05hr 95% CI 6.37-7.71) compared to periods of symptomatic remission (3.02 95% CI 2.31-3.91, p=0.042; -1.26 95% CI -1.35- -1.17, p<0.001; 4.81 95% CI 4.48–5.17, p<0.001) (**Figure 3**) (**Supplementary Tables 3 and 4**).

**Figure 3.**
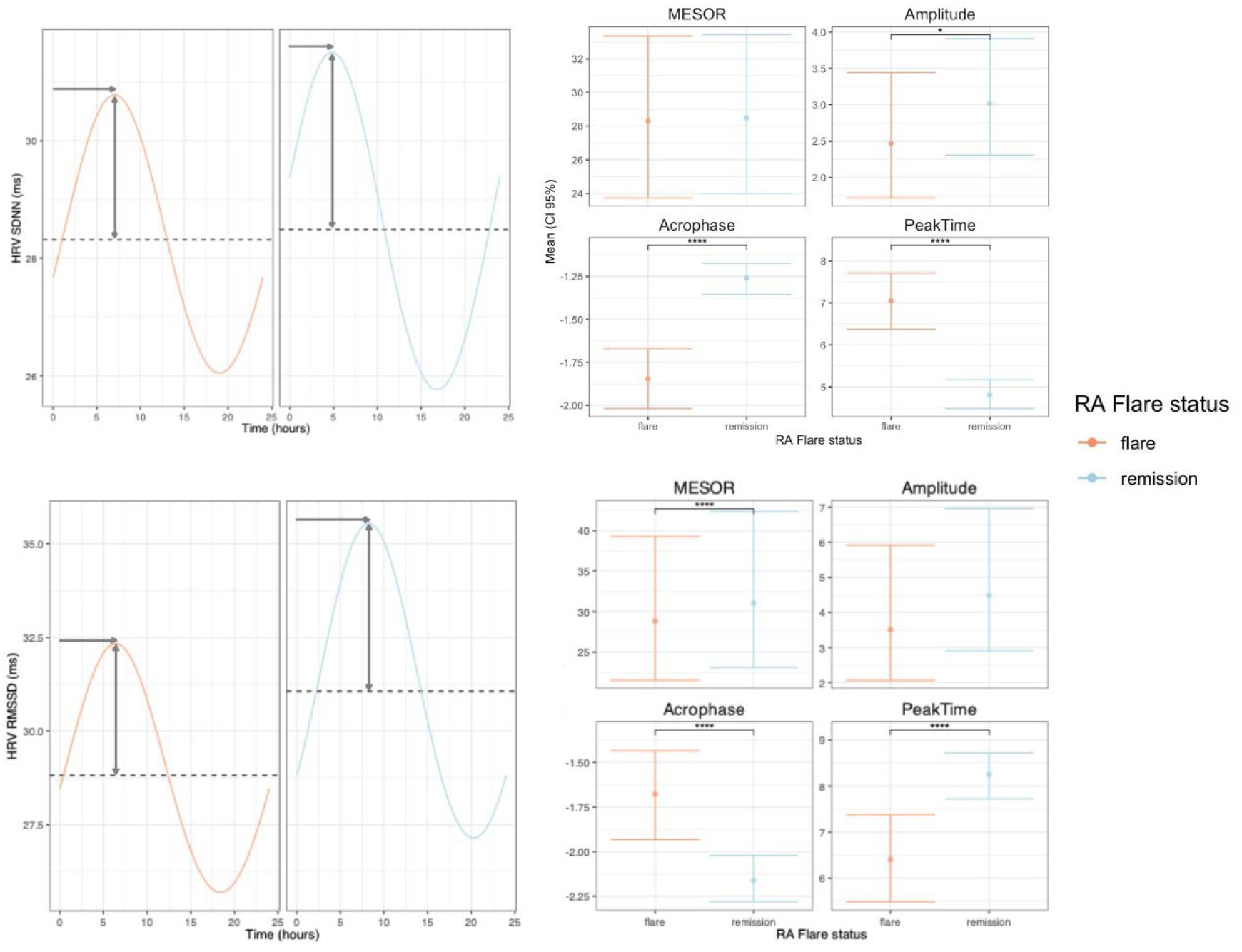
Comparison of circadian parameters of heart rate variability between periods of symptomatic flare with symptomatic remission. Stars indicate significant differences, with (+: p < 0.1; *: p < 0.05; **: p < 0.01; ***: p < 0.001, ****: p<0.0001).

### Changes Preceding Inflammatory and Symptomatic Flares

Changes in the physiological metrics RHR, daily HR, daytime HR, nighttime HR, steps and HRV (RMSSD) were evaluated on the same day as the flare, as well as 7, 14, 21, and 28 days prior to inflammatory and symptomatic flares. F1 scores balance precision and recall and are ideal when evaluating imbalanced datasets. When evaluating a model including all physiological metrics high F1 scores were maintained throughout the 28-day period prior to inflammatory flares (28 days before flare; AUC 1.00 [0.99-1.00]; F1 0.95; AUPRC 0.98; Sensitivity 0.97; Specificity 0.97) (**Figure 4a**).

**Figure 4.**
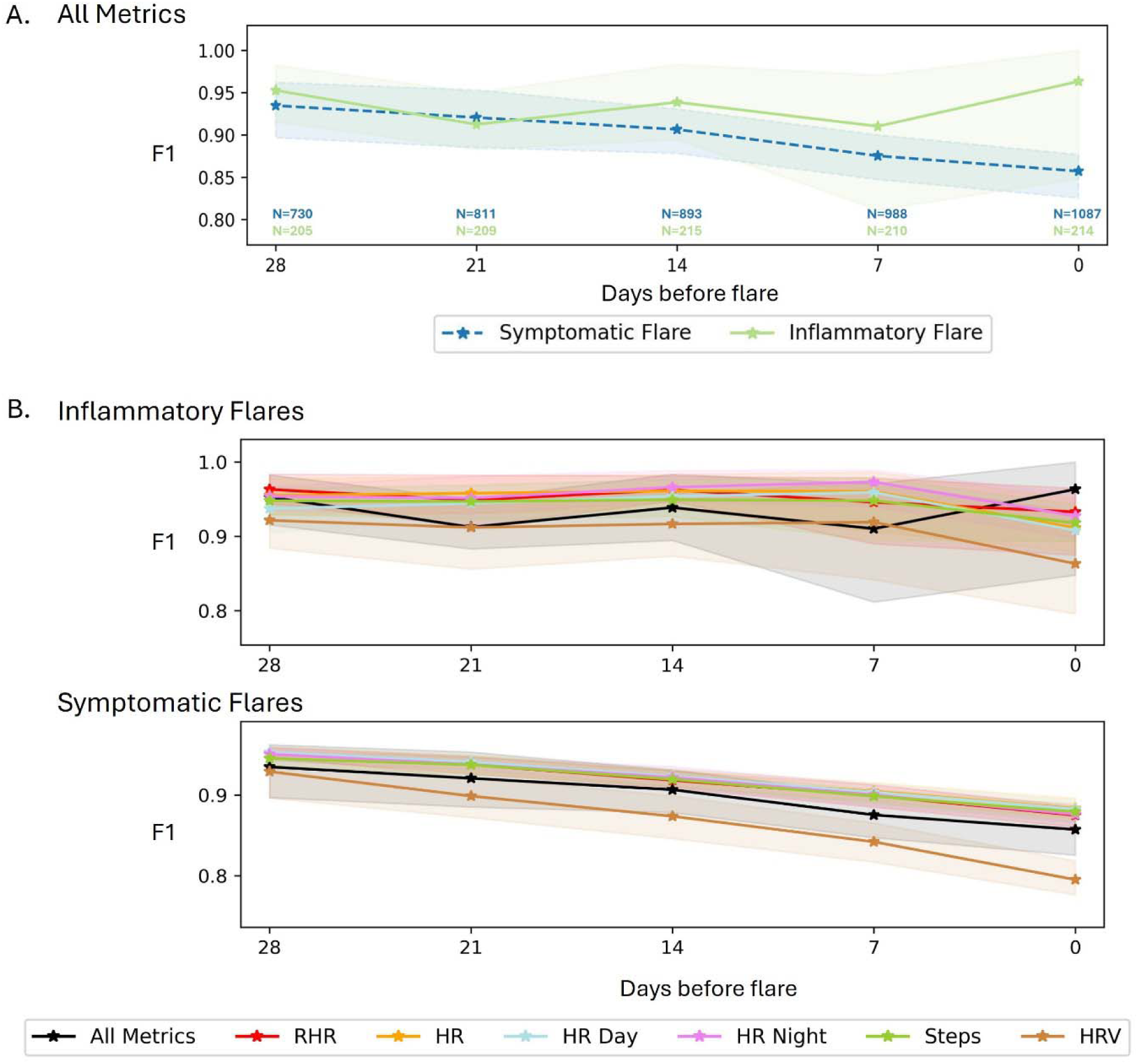
Change in physiological metrics preceding flare. (A) The association between models that include all physiological metrics, taken 7, 14, 21, and 28 days prior to the development of inflammatory and symptomatic flares, with the development of those respective flares. F1 scores summarize the performance of a model including all reported physiological metrics at each time point preceding flare development. (B) The F1 score at 7, 14, 21, and 28 days prior to inflammatory and symptomatic flare for each physiological metric measured by wearable devices.

Similar findings were found when evaluating symptomatic flares. Throughout the 28 days prior to symptomatic flares high F1 scores were observed (28 days before flare; AUC 0.99 [0.99-1.00]; F1 0.93; AUPRC 0.97; Sensitivity 0.96; Specificity 0.97).

Individual physiological metrics similarly resulted in high F1 scores throughout the 28-day period prior to inflammatory and symptomatic flares (**Figure 4b**). The AUC, AUPRC, sensitivity, specificity, and other statistical metrics evaluating the strength of association between physiological changes and subsequent inflammatory and symptomatic flares are detailed in **Supplementary Tables 5 and 6**.

## Discussion

The RA Forecast study highlights the feasibility of leveraging commercially available and widely used wearable devices to identify and potentially predict both symptomatic and inflammatory RA flares. To our knowledge, this is the first study to demonstrate that multiple physiological metrics collected from wearable devices can effectively identify these two flare types. Moreover, we provide novel evidence that these metrics exhibit significant changes several weeks prior to the onset of either flare type.

There are currently limited modalities available to monitor RA and no available means of predicting the future development of flares. Current monitoring assessments are limited to inconvenient cross-sectional assessments that often require visiting a medical facility to be performed. Furthermore, many of these assessments are performed once symptoms develop and flares have already started to occur. However, advancements in digital and wearable technology now enable the frequent passive monitoring of physiological metrics remotely and in the comfort of a user’s home.

Studies investigating the remote monitoring of RA activity using wearable sensors have predominantly focused on assessing physical activity through actigraphy, linking it to symptomatic RA activity.^8–10^ While tracking RA-related symptoms is important given their association with active inflammatory disease and their significant impact on quality of life, it is equally important to detect ongoing inflammation, which drives progressive joint damage. Although physical activity monitoring holds promise, its accuracy can be compromised by device non-compliance, resulting in incomplete capture of activity or inactivity periods. Moreover, the discordance between symptoms and inflammation in RA,^15^ underscores the need for approaches that can effectively monitor both dimensions of disease activity. RA is characterized by autonomic nervous system dysfunction, evidenced by increased HR, RHR and reduced HRV indices, reflecting reduced parasympathetic nervous system activity.^16^ These metrics can be measured by wearable devices enabling the prospect of RA activity tracking through physiologically grounded assessments. Our group has previously applied a similar approach, demonstrating that changes in such physiological metrics enable prediction of infectious states such as COVID-19^17^ and chronic inflammatory diseases such as inflammatory bowel disease.^18^

In this study, we observed significant changes in physiological metrics that were longitudinally collected via wearable devices, enabling the detection of both symptomatic and inflammatory flares. Consistent with prior research, we found significantly reduced step counts during symptomatic flares. Additionally, we identified significantly elevated mean daily and nightly HR and alterations in circadian features of HRV during symptomatic flares. Symptomatic activity may be secondary to non-inflammatory complications of RA, such as osteoarthritis or sensory nerve dependent pain. The relationship between wearable metrics and symptoms may provide a novel way to objectively evaluate and correlate patient symptoms with physiological parameters. When distinguishing periods of inflammatory flares from remission, all physiological based metrics exhibited significant differences. However, step count did not effectively differentiate between inflammation and remission. This may potentially be due to the absence of symptomatic activity in these patients, which may have preserved physical activity levels. Additionally, it may be due to a localization of inflammation to the upper extremities which may not impact mobility.

These findings underscore the potential of wearable devices to detect both symptomatic and inflammatory flares. The ability to identify and predict both inflammatory and symptomatic flares is particularly important in chronic inflammatory conditions such as RA, where inflammation, which can result in joint damage, and symptoms, which impacts quality of life, can be discordant. Importantly, we also observed that physiological metrics were altered up to four weeks before the onset of symptomatic and inflammatory flares, suggesting that there are measurable changes in the body prior to flare development. Our findings are likely detecting shifts in autonomic parameters associated with subclinical symptoms or inflammation. While further research is warranted, these results highlight the potential of wearable devices to be used for the early prediction of flares, which may enable more timely interventions. Further work utilizing machine and deep learning is needed to bring these results to the individual level and determine their predictive ability for flare.

While wearable devices hold promise for detecting and predicting inflammatory and symptomatic flares in RA, their real-world implementation presents several practical challenges that warrant further investigation. Adherence to consistent device use can vary over time and may be influenced by factors such as comfort, user engagement, and digital literacy. Further research is needed to identify which patient populations are most likely to benefit and at what points in the disease course wearable monitoring should be deployed. Integrating continuous physiological data into clinical care also requires robust infrastructure for data ingestion, storage, analysis, and visualization, while maintaining privacy and security standards. It remains unclear how best to incorporate these data into clinical decision-making, whether results should be shared with clinicians, patients, or both, and how often data should be reviewed. Furthermore, an understanding of how to respond to physiologic signals in a clinically meaningful way without contributing to alarm fatigue or unnecessary anxiety is needed. Finally, translating wearable-derived trends into actionable insights will require well-calibrated predictive models and thoughtfully designed interfaces to ensure usability and trust. Addressing these challenges is essential for advancing the integration of wearables into precision rheumatology.

Our study has several limitations. First, inflammatory assessments were conducted as part of routine clinical care rather than in a standardized protocol, which limited their frequency and our ability to precisely determine transitions between inflammation and remission. To address this, we imputed laboratory values within a +/-7-day window around each blood draw to define periods of inflammatory flare. This approach was necessary due to the variability in the timing and frequency of laboratory assessments. While this imputation strategy was intended to better approximate the duration of underlying inflammation and reduce misclassification due to isolated measurements, it may have introduced bias if CRP elevations were transient or closely linked to clinical visits that triggered therapeutic interventions. As such, this method may not fully capture the true onset or resolution dynamics of inflammatory flares. Future studies with more frequent, protocolized biomarker assessments are needed to more precisely define inflammatory states and validate wearable-based physiologic changes in relation to flare timing. Second, physiological metrics such as HRV are influenced by several physiological processes. While we accounted for common covariates such as age, sex, and BMI, other unmeasured factors may have influenced these physiological metrics.

Additionally, there are no standardized criteria for defining symptomatic flares of RA using daily symptom-based assessments. To mitigate this, we required symptoms to exhibit some chronicity, thereby increasing the likelihood that reported symptoms reflected underlying RA activity. Despite performing a sensitivity analyses, misclassification of symptomatic periods remains a possibility. A further limitation of our study is the inability to analyze the intersection of inflammatory and symptomatic activity. Only 6 participants reported both symptomatic and inflammatory flares with overlapping wearables data. Furthermore, there were only 3 participants with inflammatory flare and symptomatic remission, 0 participants with symptomatic flare and inflammatory remission and 14 participants with both inflammatory and symptomatic remission. Given the discordance between patient-reported symptoms and objective inflammatory activity in RA, future studies will be essential to evaluate how different combinations of symptomatic and inflammatory activity influence physiological signals. Another limitation is the inability to fully account for the effects of medication use, including steroids, in our analyses. Participants were on a range of therapies that varied in type, dosage, administration frequency, and timing of initiation or modification.

Additionally, the alignment between medication changes and the timing of symptom and inflammation assessments was inconsistent. These factors limit our ability to adjust for medication-related influences on the observed physiologic and clinical outcomes, as well as their impact on inflammatory and symptomatic flares. Future studies controlling for medication use will be critical to disentangling the effects of therapeutic interventions from intrinsic disease activity when interpreting wearable-derived physiological metrics. Additionally, although sleep data were collected from the wearable devices, this was not included in the current analysis. Given the complexity of these metrics and their potential relevance to RA flares, a more detailed, dedicated analysis will be carried out. A further limitation is that three different device types were used in the study with varying wear positions, sampling frequencies, and data processing algorithms. To account for these device-specific differences in our analyses, we included device type as a covariate in all multivariate models and incorporated a random intercept for each individual to account for within-person variation. Because each participant contributed data from only one device (for two participants with multiple devices, the device with more wear time was selected), this modeling approach adjusts for between-device variability while preserving the longitudinal structure of the data. Most importantly, our primary analyses focus on within-subject changes over time, rather than absolute between-subject comparisons. As such, the stability and reproducibility of a single device’s measurements over time, rather than its relationship to the ground truth, is most critical to the validity of our findings. Lastly, our dataset is imbalanced, with remission periods outnumbering flare periods. To analyze the changes in physiological metrics preceding flares, we employed mixed-effects logistic regression models, which are ideal for datasets with repeated measures and individuals contributing multiple flare events. However, this approach may bias AUC results. Consistent with prior studies involving wearable devices, we therefore focused on the F1 score, a metric better suited to evaluate performance in imbalanced datasets. No automated feature selection method was applied as our goal was to evaluate the combined association of all physiologically relevant variables collected by the devices. Advanced models incorporating feature selection and other machine learning methods are currently under development and will be presented in a separate manuscript.

### Conclusions

In conclusion, this study is the first to demonstrate that multiple longitudinal physiological metrics collected from wearable devices can effectively distinguish periods of inflammation and symptoms in RA. Moreover, we identified significant changes in these metrics occurring several weeks before the onset of symptomatic and inflammatory flares. These findings highlight the potential of wearable devices to be used for the monitoring of RA activity, for predicting flares, and in facilitating longitudinal disease management.

## Methods

### Study Population and Procedures

The RA Forecast Study is a prospective cohort study that enrolled individuals with RA from throughout the United States between February 2023 and February 2024. Participants were recruited through study flyers and an email advertisement to individuals with RA who visited a rheumatology clinic in the Mount Sinai Health System. Inclusion criteria included (1) a diagnosis with RA, (2) age ≥18 years, (3) use of a medication to treat RA, and (4) the willingness to use a wearable device. Exclusion criteria included pregnancy, the presence of other chronic medical conditions, a pacemaker or defibrillator, or the use of medications that impact HR or HRV (ie. beta blockers, benzodiazepines). Participants could use their own wearable device (Apple Watch, Fitbit, or Oura Ring). If a participant did not have a wearable device an Apple Watch or Fitbit would be loaned for the duration of study participation. There was no compensation provided to participants.

Participants downloaded ehive^19^, our custom app, to their smartphone. Within ehive they verified inclusion criteria, signed electronic consent and linked their wearable device. Participants provided baseline demographic information and their medical history. Participants answered daily patient reported outcomes surveys in ehive and uploaded any C-reactive protein (CRP) laboratory test results collected as part of their standard of care management. Participants were free to provide as much or as little data as they would like, with smart phone push notifications sent to those who contributed wearable or survey data fewer than four days per week. It was recommended that wearable devices be used for a minimum of 8 hours per day and at least 4 days per week. Custom smart phone push notifications and emails were used as light touch engagement measures to maintain compliance.

### Wearable Devices

The study utilized the Apple Watch, Fitbit, and Oura Ring, which are three commercially available wearable devices. Both the Apple Watch and Fitibit devices are wrist worn, while the Oura Ring is sized and fit on an individual’s finger. Heart rate (HR), resting heart rate (RHR), HRV, and steps were collected from the three devices. All three devices contain photoplethysmography optical sensors that assess capillary volume changes. These enable the sensing of heart beats and the time difference between each heartbeat, or the interbeat intervals (IBI), for the calculation of HRV.^20,21^ Each device contains an actigraphy sensor tracking step counts. HR is reported as beats per minute (BPM) and RHR is defined as an estimation of participants lowest HR during rest periods.

All three devices calculate HRV. The Apple Watch calculates SDNN. This is a time domain HRV metric that reflects both sympathetic and parasympathetic nervous system activity. The Fitbit and Oura Ring calculate HRV as RMSSD. This time domain metric is influenced more by the parasympathetic nervous system compared to the sympathetic nervous system, with lower values reflecting increased sympathetic tone.^22^ IBI data was also collected from the Apple Watch of participants, enabling calculation of HRV in the RMSSD domain and the combination of this data with that collected from the Fitbit and Oura Ring.

The commercially available physiological data collected from the Apple Watch, Fitbit, and Oura Ring are pre-processed by each manufacturer using proprietary algorithms designed to minimize artifacts and signal noise. The Apple Watch reports HR at variable sampling intervals, with frequencies reaching up to one reading per second in our cohort. HRV (RMSSD) is derived from IBI segments of approximately one-minute duration, with an average of six HRV readings per participant per day. In comparison, in our cohort, the Fitbit and Oura Ring provided HR data at fixed intervals of one minute and five minutes, respectively, and measured HRV every five minutes during sleep periods. To support analysis across varying time scales, these features were aggregated to generate hourly, daytime, nighttime, and full 24-hour daily summary metrics.

### Laboratory and Clinical Assessments

RA flares were defined as (1) inflammatory flares, denoting the presence of underlying inflammation, and (2) symptomatic flares, characterized by the presence of symptoms. Laboratory measures of inflammation were collected as part of each participant’s standard of care management throughout the study period. An inflammatory flare was defined by a CRP value >5 mg/dL. Given that inflamed and non-inflamed states are present beyond the point in time that a laboratory measure is drawn, and to avoid misclassification of inflammatory status around the laboratory test, the CRP values were imputed in a +/-7-day window around the date of each blood draw.^23^

Symptomatic activity was characterized using the Routine Assessment of Patient Index Data (RAPID-3) scale.^4^ This survey includes a subset of the primary variables found in the Multi-dimensional HAQ (MD-HAQ) and is designed for self-administration. The RAPID-3 includes an assessment of physical function, a patient global assessment for pain, and a patient global health assessment. It has been validated and shown to correlate with DAS28 and CDAI scores in RA.^24,25^ It is scored from 0-30, with a score >3 consistent with active RA of mild, moderate or high disease severity; score ≤3 is classified as remission.^4^ Most symptom assessments are collected on a less than daily frequency. Thus, there is no consensus on the number of daily symptom-based surveys meeting criteria for symptomatic disease activity that are required to classify a period as a symptom flare. To evaluate several criteria, we performed a sensitivity analysis for several definitions of symptomatic flare in our study (**Supplementary Figure** 3). Based on this analysis, we defined symptomatic flares as having at least 4 RAPID-3 surveys answered in a 7-day period, with at least 3 of these surveys meeting criteria (RAPID-3 >3) for symptomatic activity. Each 7-day period was analyzed as a rolling window that includes daily survey data that extends 3 days before and 3 days after each day being analyzed.

### Statistical Analysis

Physiological metrics were analyzed using linear mixed-effect (LME) models, to quantify differences between flare and remission periods. This and the following statistical models included fixed effects for the covariates of age, sex, device type and body mass index (BMI) and a random intercept for each participant. This allows each individual to have their own baseline value, thereby accounting for within-subject correlation, along with population-level influence of age, sex, BMI and signal-quality differences across devices. Missing values for age (n=5) and sex (n=1) were imputed using multivariate imputation that uses the entire dataset of available features including demographics and device features across all participants. Flare and remission states were incorporated as predictors in each model. The models were fitted on a complete-case dataset that analyzed only days for which physiological variables were present, with days with missing values excluded. The “*lme*” function from the “*nlme*” R package was used to fit all physiological metrics.^26^ The *emmeans* package was used to estimate marginal means with 95% confidence intervals (CI).^27^ HR was analyzed as a mean value over three time periods: (1) daily: assessed over the entire 24 hour period, (2) daytime: including data from 8:00am-10:00pm, (3) nighttime: including data from 10:00pm-8:00am. Below shows LME model equation for RHR, similarly performed for all physiological metrics:

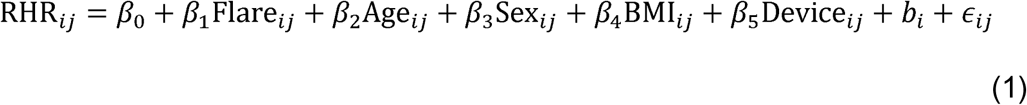

with

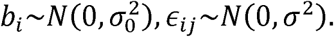

*RHR_ij_* is resting heart rate at day *j* of participant, *i*, *β*_1_ captures mean difference in RHR between flare and remission days after adjusting for age, sex, BMI, and device type (*β*_2_ – *β*_5) ._ *b_i_* is the random intercept, modeling within-subject correlation and allowing each participant to have an individual baseline RHR. Our linear mixed-effects models yield reliable estimates under the assumption that missing wearable data are missing at random (MAR), an assumption considered appropriate in this context. Missingness may arise from device-related technical issues or variable participant engagement; however, these effects are mitigated through inclusion of relevant covariates, temporal alignment of wearable data with clinical assessments, and the use of within-subject comparisons across a large longitudinal dataset. To assess whether missing data were associated with disease activity, we performed a paired t-test comparing the percentage of missing days during flare and remission periods on a per-participant basis. For both symptomatic and inflammatory flare categories, the p-values were 0.49 and 0.23, respectively, indicating no statistically significant difference in missingness between flare and remission states. These findings support the assumption that missing data is missing at random (MAR), rather than being systematically related to disease activity.

HRV has a daily circadian pattern over a 24-hour period that can be modelled using a non-linear Cosinor model.^28^ These dynamic changes are therefore not accounted for using traditional means of assessment such as averages or a range of values. Additionally, wearable devices collect HRV metrics in a non-uniform pattern. To model such longitudinal features methods that assess circadian patterns are needed.

COSINOR methods model circadian rhythms and have been applied to the evaluation of HRV.^29,30^

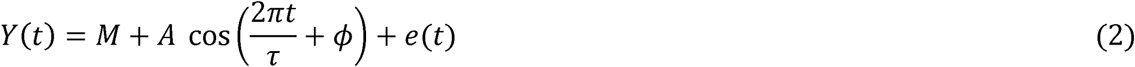

The circadian pattern can be described where M, A and ϕ describe the cyclic parameters (1) MESOR (M): a rhythm-adjusted mean of the circadian pattern; (2) Amplitude (A): difference between the peak and the mean value giving a measure of half the extent of variation within a day; and (3) Acrophase (ϕ): a measure of the time when peak values occur each day. Acrophase (ϕ) is related to peak time (*t_p_*) as 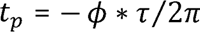. Our team has extended this work to include a mixed-effect model framework enabling evaluation of changes in daily circadian patterns of HRV through Cosinor mixed effects models. This analysis was carried out using the CosinorRmixedeffects R package,^31^ which models HRV circadian rhythm over 24 hours, with τ hours, allowing robust handling of missing hourly HRV samples across the 24 hours. To improve model fit and ensure the assumptions of normality and homoscedasticity were met, a log transformation was applied to the HRV RMSSD and SDNN values prior to modeling with the cosinor mixed-effects framework. Model estimates were subsequently transformed back to the original scale for interpretation. This transformation resulted in improved model diagnostics, including residuals that approximated a normal distribution with constant variance. Goodness-of-fit was further supported by residual plots, which are presented in **Supplementary Figure 4 and Supplementary Figure 5**, for both inflammatory and symptomatic flare models. A mixed-effects logistic regression (LR) model was employed to evaluate the temporal association of the individual physiological metrics, and their combination, in determining the probability of having an inflammatory or symptomatic flare. Predictions were derived on the same day, 7, 14, 21, and 28 days prior to flares. A complete-case analysis was performed by only using days when all features are available. Model performance was estimated using the area under the curve (AUC), area under the precision-recall curve (AUPRC), threshold, sensitivity, specificity, F1, precision, recall, and accuracy. The “*glmer”* function from the “*lme4*” R package was used for mixed-effects LR.^32^ This approach incorporated fixed effects for age, sex, BMI, and device type, as well as a random intercept for each participant to account for within-subject correlation across repeated measures. The included physiological metrics were RHR, daily HR, daytime HR, nighttime HR, step count, and HRV (RMSSD).

### Study Approval

This study was approved by the Mount Sinai Institutional Review Board. Written informed consent was obtained from all participants prior to participation. All research was carried out in accordance with relevant guidelines and regulations. Research was performed in accordance with the Declaration of Helsinki.

## Author contributions

PS, MD, JKW, KL, DH, BES, MSF, PG, RPH made substantial contributions to the conception or design of the work, or the acquisition, analysis or interpretation of the data for the work; made contributions to the drafting of the work or revising it critically for important intellectual content; provided final approval of the version to be published and agree to be accountable for all aspects of the work in ensuring that questions related to the accuracy or integrity of any part of the work are addressed.

## Data Availability

The datasets analyzed during the current study is available from the corresponding author on reasonable request.

