## Supplemental Material for "Wearable Devices Detect Physiological Changes that Precede and Are Associated with Symptomatic and Inflammatory Rheumatoid Arthritis Flares"

Supplemental Table 1. Additional study characteristics relating to study duration, device compliance, survey and C-reactive protein laboratory records, flare prevalence, and missing data during flares and remission. The mean and standard deviations (SD) are across all participants and N shows count across the entire cohort.

|  | Overall Cohort | Apple Watch | Fitbit | Oura Ring |
| --- | --- | --- | --- | --- |
| Study duration: | | | | |
| Number of Days of follow-up from enrollment to study withdrawal/termination, mean (SD) | 246.85  (63.39) | 252.76  (53.00) | 236.63  (86.16) | 234.33  (28.01) |
| Number of Days between first and last day with any RAPID3 survey/CRP/wearables data (data days), mean (SD) | 189.42  (95.36) | 191.00  (99.96) | 179.31  (95.16) | 225.33  (29.02) |
| Number of Days between first and last day with any RAPID3 survey/CRP/wearables data (data days), N | 10,039 | 6,494 | 2,869 | 676 |
| Device compliance: | | | | |
| Number of Days with data from wearable devices, mean (SD) | 154.40  (93.23) | 165.94  (96.33) | 126.31  (88.86) | 173.33  (67.49) |
| Number of Days with data from wearable devices, N | 8,183 | 5,642 | 2,021 | 520 |
| Number of Hours with data from wearable devices, mean (SD) | 2,697.83  (2,004.10) | 2,767.44  (1,988.46) | 2,462.38  (2,084.21) | 3,164.67  (2,398.65) |
| Number of Hours with data from wearable devices, N | 142,985 | 94,093 | 39,398 | 9,494 |
| Mean number of Hours per Day with data from wearable devices, mean (SD) | 16.38  (4.60) | 15.61  (4.06) | 18.03  (5.01) | 16.36  (7.60) |
| Survey and lab records: | | | | |
| Number of Days where RAPID3 daily survey was answered, mean (SD) | 72.96  (72.35) | 84.12  (82.23) | 57.88  (47.67) | 27.00  (19.7) |
| Number of Days where RAPID3 daily survey was answered, N | 3,867 | 2,860 | 926 | 81 |
| Percentage of Days out of data days where RAPID3 daily survey was answered, mean (SD) % | 37.15  (25.73) | 41.64  (27.5) | 32.43  (20.51) | 11.36  (7.91) |
| Number of Days in the study period with CRP lab, mean (SD) | 1.28  (1.39) | 1.26  (1.48) | 1.25  (1.24) | 1.67  (1.53) |
| Number of Days in the study period with CRP lab, N | 68 | 43 | 20 | 5 |
| Percentage of Days out of data days with CRP lab, mean (SD) % | 0.74  (0.74) | 0.61  (0.68) | 1.02  (0.84) | 0.69  (0.63) |
| Flare assessment: | | | | |
| Number of Days in the study period with symptomatic flare, mean (SD) | 37.30  (64.14) | 42.00  (74.75) | 32.69  (41.34) | 8.67  (12.5) |
| Number of Days in the study period with symptomatic flare, N | 1,977 | 1,428 | 523 | 26 |
| Percentage of Days out of data days with symptomatic flare, mean (SD) % | 18.14  (26.05) | 18.01  (26.97) | 21.14  (26.4) | 3.65  (5.03) |
| Number of Days in the study period with inflammatory flare, mean (SD) | 5.09  (10.17) | 3.97  (9.28) | 5.63  (7.50) | 15.00  (25.98) |
| Number of Days in the study period with inflammatory flare, N | 270 | 135 | 90 | 45 |
| Percentage of Days out of data days with inflammatory flare, mean (SD) % | 4.00  (8.77) | 3.00  (7.69) | 5.70  (10.73) | 6.12  (10.6) |
| Missing wearable assessment: | | | | |
| Number of Days with missing wearable data during symptomatic flare, N | 42 | 42 | 0 | 0 |
| Percentage of Days with missing wearable data out of days with symptomatic flare, mean (SD) % | 1.22  (3.57) | 1.99  (4.43) | 0  (0) | 0  (0) |
| Number of Days with missing wearable data during symptomatic remission, N | 47 | 30 | 13 | 4 |
| Percentage of Days with missing wearable data out of days with symptomatic remission, mean (SD) % | 5.04  (18.57) | 1.78 (3.82) | 12.51  (32.94) | 1.17  (2.03) |
| Number of Days with missing wearable data during inflammatory flare, N | 55 | 35 | 13 | 7 |
| Percentage of Days with missing wearable data out of days with inflammatory flare, mean (SD) % | 14.44  (28.27) | 14.29  (26.23) | 14.44  (35.38) | 15.56  (NA) |
| Number of Days with missing wearable data during inflammatory remission, N | 71 | 37 | 24 | 10 |
| Percentage of Days with missing wearable data out of days with inflammatory remission, mean (SD) % | 8.61  (17.66) | 6.06  (10.76) | 10.65  (25.09) | 20.00  (28.28) |

Supplemental Table 2. Linear mixed effect model results for each physiological metric during both symptomatic and inflammatory flares. Results show flare vs remission contrasts based on expected marginal means, standard error and associated p-values.

| **Flare** | **Physiological Metric** | **Flare-Remission** | **Standard Error** | **P-Value** |
| --- | --- | --- | --- | --- |
| Symptomatic Flare | RHR | -0.12 | 0.27 | 0.66 |
|  | HR | 1.26 | 0.60 | 0.04 |
|  | Steps | -515.52 | 177.47 | 0.004 |
|  | HR Day | 0.50 | 0.65 | 0.44 |
|  | HR Night | 2.90 | 0.70 | <0.0001 |
| Inflammatory Flare | RHR | 4.97 | 0.84 | <0.0001 |
|  | HR | 5.99 | 1.66 | <0.001 |
|  | Steps | -992.53 | 583.02 | 0.09 |
|  | HR Day | 6.44 | 1.65 | <0.001 |
|  | HR Night | 5.47 | 1.86 | <0.004 |

Supplemental Table 3. Logarithmically transformed HRV RMSSD cosinor model results for symptomatic and inflammatory flares, across all devices. For each metric of mesor, amplitude, acrophase and peak time, the results show flare vs remission contrasts and associated p-values. Shown differences and P-values are in log scale.

| **Flare** | **Metric** | **Flare-Remission** | **Standard Error** | **P-Value** |
| --- | --- | --- | --- | --- |
| Symptomatic Flare | Mesor | -0.033 | 0.0066 | <0.001 |
|  | Amplitude | -0.009 | 0.0064 | 0.17 |
|  | Acrophase | 0.482 | 0.1348 | 0.002 |
|  | Peak Time | -1.843 | 0.5150 | 0.002 |
| Inflammatory Flare | Mesor | -0.126 | 0.0180 | <0.001 |
|  | Amplitude | -0.031 | 0.0150 | 0.04 |
|  | Acrophase | -0.023 | 0.9870 | 0.98 |
|  | Peak Time | 0.087 | 3.7702 | 0.98 |

Supplemental Table 4. Logarithmically transformed HRV SDNN cosinor model results for symptomatic and inflammatory flares, only available for Apple watch. For each metric of mesor, amplitude, acrophase and peak time, the results show flare vs remission contrasts and associated p-values. Shown differences and P-values are in log scale.

| **Flare** | **Metric** | **Flare-Remission** | **Standard Error** | **P-Value** |
| --- | --- | --- | --- | --- |
| Symptomatic Flare | Mesor | -0.003 | 0.005 | 0.62 |
|  | Amplitude | -0.007 | 0.004 | 0.04 |
|  | Acrophase | -0.586 | 0.101 | <0.001 |
|  | Peak Time | 2.240 | 0.385 | <0.001 |
| Inflammatory Flare | Mesor | -0.021 | 0.025 | 0.39 |
|  | Amplitude | 0.040 | 0.014 | 0.006 |
|  | Acrophase | -0.254 | 0.281 | 0.35 |
|  | Peak Time | 0.971 | 1.074 | 0.35 |

Supplemental Table 5. The ability of physiological parameters to identify inflammatory flares up to 28 days prior to the event. The full model includes daily heart rate, steps, resting heart rate, daytime heart rate, nighttime heart rate, and heart rate variability. N represents the number of days in the logistic regression model.

| Number of Days Prior To Flare | Predictor | AUC (95% CI) | AUPRC | Threshold | Sensitivity | Specificity | F1 | Precision | Recall | Accuracy | N |
| --- | --- | --- | --- | --- | --- | --- | --- | --- | --- | --- | --- |
| Day of the Flare | Full Model | 1.00 (0.99-1.00) | 0.96 | 0.4 | 0.98 | 0.99 | 0.96 | 0.95 | 0.98 | 0.99 | 214 |
| Day of the Flare | RHR | 0.99 (0.99-1.00) | 0.97 | 0.25 | 0.98 | 0.97 | 0.93 | 0.89 | 0.98 | 0.97 | 702 |
| Day of the Flare | Daily HR | 0.99 (0.99-1.00) | 0.96 | 0.24 | 0.99 | 0.95 | 0.91 | 0.84 | 0.99 | 0.96 | 769 |
| Day of the Flare | Daytime HR | 0.99 (0.99-0.99) | 0.96 | 0.26 | 0.99 | 0.95 | 0.91 | 0.84 | 0.99 | 0.96 | 753 |
| Day of the Flare | Nighttime HR | 0.99 (0.99-1.00) | 0.97 | 0.24 | 0.99 | 0.95 | 0.93 | 0.87 | 0.99 | 0.96 | 661 |
| Day of the Flare | Daily Steps | 0.99 (0.98-0.99) | 0.95 | 0.29 | 0.99 | 0.96 | 0.92 | 0.86 | 0.99 | 0.96 | 753 |
| Day of the Flare | HRV | 0.97 (0.94-0.99) | 0.93 | 0.31 | 0.92 | 0.93 | 0.86 | 0.82 | 0.92 | 0.93 | 297 |
| 7 Days Before Flare | Full Model | 0.99 (0.97-1.00) | 0.95 | 0.32 | 0.94 | 0.96 | 0.91 | 0.9 | 0.94 | 0.95 | 210 |
| 7 Days Before Flare | RHR | 1.00 (0.99-1.00) | 0.99 | 0.3 | 0.96 | 0.97 | 0.95 | 0.93 | 0.96 | 0.97 | 686 |
| 7 Days Before Flare | Daily HR | 1.00 (1.00-1.00) | 0.99 | 0.32 | 0.97 | 0.98 | 0.96 | 0.95 | 0.97 | 0.98 | 740 |
| 7 Days Before Flare | Daytime HR | 1.00 (1.00-1.00) | 0.99 | 0.32 | 0.97 | 0.98 | 0.96 | 0.95 | 0.97 | 0.98 | 723 |
| 7 Days Before Flare | Nighttime HR | 1.00 (0.99-1.00) | 0.99 | 0.36 | 0.96 | 0.99 | 0.97 | 0.99 | 0.96 | 0.98 | 640 |
| 7 Days Before Flare | Daily Steps | 1.00 (0.99-1.00) | 0.98 | 0.32 | 0.97 | 0.98 | 0.95 | 0.93 | 0.97 | 0.97 | 734 |
| 7 Days Before Flare | HRV | 0.98 (0.97-0.99) | 0.96 | 0.32 | 0.91 | 0.97 | 0.92 | 0.94 | 0.91 | 0.95 | 284 |
| 14 Days Before Flare | Full Model | 0.99 (0.99-1.00) | 0.97 | 0.32 | 0.98 | 0.96 | 0.94 | 0.9 | 0.98 | 0.96 | 215 |
| 14 Days Before Flare | RHR | 1.00 (1.00-1.00) | 0.99 | 0.31 | 0.99 | 0.97 | 0.96 | 0.93 | 0.99 | 0.98 | 670 |
| 14 Days Before Flare | Daily HR | 1.00 (1.00-1.00) | 0.99 | 0.3 | 0.98 | 0.98 | 0.96 | 0.94 | 0.98 | 0.98 | 710 |
| 14 Days Before Flare | Daytime HR | 1.00 (1.00-1.00) | 0.99 | 0.29 | 0.98 | 0.97 | 0.95 | 0.93 | 0.98 | 0.97 | 696 |
| 14 Days Before Flare | Nighttime HR | 1.00 (1.00-1.00) | 0.99 | 0.33 | 0.98 | 0.98 | 0.97 | 0.95 | 0.98 | 0.98 | 628 |
| 14 Days Before Flare | Daily Steps | 1.00 (1.00-1.00) | 0.99 | 0.28 | 0.99 | 0.97 | 0.95 | 0.91 | 0.99 | 0.97 | 702 |
| 14 Days Before Flare | HRV | 0.99 (0.98-0.99) | 0.97 | 0.31 | 0.95 | 0.94 | 0.92 | 0.89 | 0.95 | 0.94 | 284 |
| 21 Days Before Flare | Full Model | 0.99 (0.99-1.00) | 0.97 | 0.32 | 0.98 | 0.96 | 0.94 | 0.9 | 0.98 | 0.96 | 215 |
| 21 Days Before Flare | RHR | 0.99 (0.98-1.00) | 0.96 | 0.31 | 0.97 | 0.93 | 0.91 | 0.87 | 0.97 | 0.94 | 209 |
| 21 Days Before Flare | Daily HR | 1.00 (1.00-1.00) | 0.99 | 0.3 | 0.98 | 0.97 | 0.95 | 0.92 | 0.98 | 0.97 | 626 |
| 21 Days Before Flare | Daytime HR | 1.00 (1.00-1.00) | 0.99 | 0.34 | 0.99 | 0.97 | 0.96 | 0.93 | 0.99 | 0.98 | 671 |
| 21 Days Before Flare | Nighttime HR | 1.00 (1.00-1.00) | 0.99 | 0.32 | 1 | 0.96 | 0.94 | 0.9 | 1 | 0.97 | 658 |
| 21 Days Before Flare | Daily Steps | 1.00 (1.00-1.00) | 0.99 | 0.31 | 0.99 | 0.96 | 0.95 | 0.91 | 0.99 | 0.97 | 596 |
| 21 Days Before Flare | HRV | 1.00 (1.00-1.00) | 0.99 | 0.3 | 0.99 | 0.96 | 0.95 | 0.9 | 0.99 | 0.97 | 659 |
| 28 Days Before Flare | Full Model | 1.00 (0.99-1.00) | 0.98 | 0.39 | 0.97 | 0.97 | 0.95 | 0.94 | 0.97 | 0.97 | 205 |
| 28 Days Before Flare | RHR | 1.00 (1.00-1.00) | 0.99 | 0.29 | 0.99 | 0.97 | 0.96 | 0.93 | 0.99 | 0.98 | 611 |
| 28 Days Before Flare | Daily HR | 1.00 (1.00-1.00) | 0.99 | 0.34 | 0.99 | 0.97 | 0.96 | 0.93 | 0.99 | 0.98 | 660 |
| 28 Days Before Flare | Daytime HR | 1.00 (0.99-1.00) | 0.99 | 0.31 | 0.99 | 0.96 | 0.94 | 0.89 | 0.99 | 0.97 | 644 |
| 28 Days Before Flare | Nighttime HR | 1.00 (1.00-1.00) | 0.99 | 0.33 | 0.99 | 0.97 | 0.95 | 0.92 | 0.99 | 0.97 | 589 |
| 28 Days Before Flare | Daily Steps | 1.00 (1.00-1.00) | 0.99 | 0.31 | 0.99 | 0.96 | 0.95 | 0.91 | 0.99 | 0.97 | 653 |
| 28 Days Before Flare | HRV | 0.99 (0.98-1.00) | 0.97 | 0.31 | 0.98 | 0.93 | 0.92 | 0.87 | 0.98 | 0.94 | 262 |

Supplemental Table 6. The ability of physiological parameters to identify symptomatic flares up to 28 days prior to the event. The full model includes daily heart rate, steps, resting heart rate, daytime heart rate, nighttime heart rate, and heart rate variability. N represents the number of days in the logistic regression model.

| Number of Days Prior To Flare | Predictor | AUC (95% CI) | AUPRC | Threshold | Sensitivity | Specificity | F1 | Precision | Recall | Accuracy | N |
| --- | --- | --- | --- | --- | --- | --- | --- | --- | --- | --- | --- |
| Day of the Flare | Full Model | 0.97  (0.97-0.98) | 0.93 | 0.25 | 0.96 | 0.9 | 0.86 | 0.78 | 0.96 | 0.92 | 1087 |
| Day of the Flare | RHR | 0.97  (0.97-0.98) | 0.96 | 0.37 | 0.9 | 0.91 | 0.87 | 0.85 | 0.9 | 0.91 | 4540 |
| Day of the Flare | Daily HR | 0.97  (0.97-0.98) | 0.96 | 0.4 | 0.89 | 0.93 | 0.88 | 0.87 | 0.89 | 0.92 | 4757 |
| Day of the Flare | Daytime HR | 0.97  (0.97-0.98) | 0.96 | 0.39 | 0.9 | 0.93 | 0.88 | 0.87 | 0.9 | 0.92 | 4685 |
| Day of the Flare | Nighttime HR | 0.97  (0.97-0.98) | 0.96 | 0.35 | 0.91 | 0.92 | 0.88 | 0.85 | 0.91 | 0.91 | 4385 |
| Day of the Flare | Daily Steps | 0.97  (0.97-0.98) | 0.96 | 0.35 | 0.9 | 0.91 | 0.88 | 0.86 | 0.9 | 0.91 | 4681 |
| Day of the Flare | HRV | 0.96  (0.95-0.97) | 0.9 | 0.19 | 0.97 | 0.84 | 0.8 | 0.68 | 0.97 | 0.87 | 1375 |
| 7 Days Before Flare | Full Model | 0.98 (0.97-0.98) | 0.95 | 0.29 | 0.96 | 0.92 | 0.88 | 0.81 | 0.96 | 0.93 | 988 |
| 7 Days Before Flare | RHR | 0.98 (0.98-0.98) | 0.97 | 0.39 | 0.92 | 0.93 | 0.9 | 0.88 | 0.92 | 0.93 | 4141 |
| 7 Days Before Flare | Daily HR | 0.98 (0.98-0.99) | 0.97 | 0.44 | 0.91 | 0.94 | 0.9 | 0.9 | 0.91 | 0.93 | 4334 |
| 7 Days Before Flare | Daytime HR | 0.98 (0.98-0.99) | 0.97 | 0.38 | 0.92 | 0.94 | 0.9 | 0.89 | 0.92 | 0.93 | 4271 |
| 7 Days Before Flare | Nighttime HR | 0.98 (0.98-0.98) | 0.97 | 0.38 | 0.92 | 0.93 | 0.9 | 0.88 | 0.92 | 0.93 | 3998 |
| 7 Days Before Flare | Daily Steps | 0.98 (0.98-0.98) | 0.97 | 0.36 | 0.93 | 0.92 | 0.9 | 0.87 | 0.93 | 0.92 | 4276 |
| 7 Days Before Flare | HRV | 0.97 (0.97-0.98) | 0.93 | 0.3 | 0.97 | 0.89 | 0.84 | 0.74 | 0.97 | 0.91 | 1228 |
| 14 Days Before Flare | Full Model | 0.99 (0.98-0.99) | 0.97 | 0.31 | 0.96 | 0.95 | 0.91 | 0.86 | 0.96 | 0.95 | 893 |
| 14 Days Before Flare | RHR | 0.99 (0.98-0.99) | 0.98 | 0.38 | 0.94 | 0.94 | 0.92 | 0.9 | 0.94 | 0.94 | 3847 |
| 14 Days Before Flare | Daily HR | 0.99 (0.98-0.99) | 0.98 | 0.4 | 0.94 | 0.94 | 0.92 | 0.91 | 0.94 | 0.94 | 4023 |
| 14 Days Before Flare | Daytime HR | 0.99 (0.98-0.99) | 0.98 | 0.37 | 0.94 | 0.95 | 0.92 | 0.91 | 0.94 | 0.94 | 3964 |
| 14 Days Before Flare | Nighttime HR | 0.99 (0.98-0.99) | 0.98 | 0.39 | 0.94 | 0.94 | 0.92 | 0.9 | 0.94 | 0.94 | 3714 |
| 14 Days Before Flare | Daily Steps | 0.99 (0.98-0.99) | 0.98 | 0.39 | 0.94 | 0.93 | 0.92 | 0.9 | 0.94 | 0.94 | 3964 |
| 14 Days Before Flare | HRV | 0.98 (0.97-0.99) | 0.95 | 0.29 | 0.97 | 0.92 | 0.87 | 0.79 | 0.97 | 0.93 | 1108 |
| 21 Days Before Flare | Full Model | 0.99 (0.98-0.99) | 0.97 | 0.34 | 0.96 | 0.96 | 0.92 | 0.88 | 0.96 | 0.96 | 811 |
| 21 Days Before Flare | RHR | 0.99 (0.99-0.99) | 0.98 | 0.4 | 0.94 | 0.96 | 0.94 | 0.93 | 0.94 | 0.95 | 3593 |
| 21 Days Before Flare | Daily HR | 0.99 (0.99-0.99) | 0.98 | 0.37 | 0.95 | 0.96 | 0.94 | 0.93 | 0.95 | 0.95 | 3771 |
| 21 Days Before Flare | Daytime HR | 0.99 (0.99-0.99) | 0.99 | 0.38 | 0.95 | 0.96 | 0.94 | 0.94 | 0.95 | 0.95 | 3714 |
| 21 Days Before Flare | Nighttime HR | 0.99 (0.99-0.99) | 0.98 | 0.41 | 0.95 | 0.95 | 0.94 | 0.92 | 0.95 | 0.95 | 3488 |
| 21 Days Before Flare | Daily Steps | 0.99 (0.99-0.99) | 0.98 | 0.39 | 0.94 | 0.96 | 0.94 | 0.93 | 0.94 | 0.95 | 3716 |
| 21 Days Before Flare | HRV | 0.98 (0.98-0.99) | 0.95 | 0.26 | 0.96 | 0.94 | 0.9 | 0.84 | 0.96 | 0.95 | 995 |
| 28 Days Before Flare | Full Model | 0.99 (0.99-1.00) | 0.97 | 0.37 | 0.96 | 0.97 | 0.93 | 0.91 | 0.96 | 0.97 | 730 |
| 28 Days Before Flare | RHR | 0.99 (0.99-0.99) | 0.99 | 0.39 | 0.95 | 0.97 | 0.95 | 0.95 | 0.95 | 0.96 | 3365 |
| 28 Days Before Flare | Daily HR | 0.99 (0.99-0.99) | 0.99 | 0.39 | 0.95 | 0.97 | 0.95 | 0.95 | 0.95 | 0.96 | 3535 |
| 28 Days Before Flare | Daytime HR | 0.99 (0.99-0.99) | 0.99 | 0.41 | 0.95 | 0.98 | 0.95 | 0.96 | 0.95 | 0.96 | 3476 |
| 28 Days Before Flare | Nighttime HR | 0.99 (0.99-0.99) | 0.99 | 0.38 | 0.96 | 0.97 | 0.95 | 0.94 | 0.96 | 0.96 | 3273 |
| 28 Days Before Flare | Daily Steps | 0.99 (0.99-0.99) | 0.99 | 0.4 | 0.95 | 0.96 | 0.95 | 0.95 | 0.95 | 0.96 | 3489 |
| 28 Days Before Flare | HRV | 0.99 (0.98-0.99) | 0.96 | 0.35 | 0.96 | 0.97 | 0.93 | 0.9 | 0.96 | 0.97 | 893 |

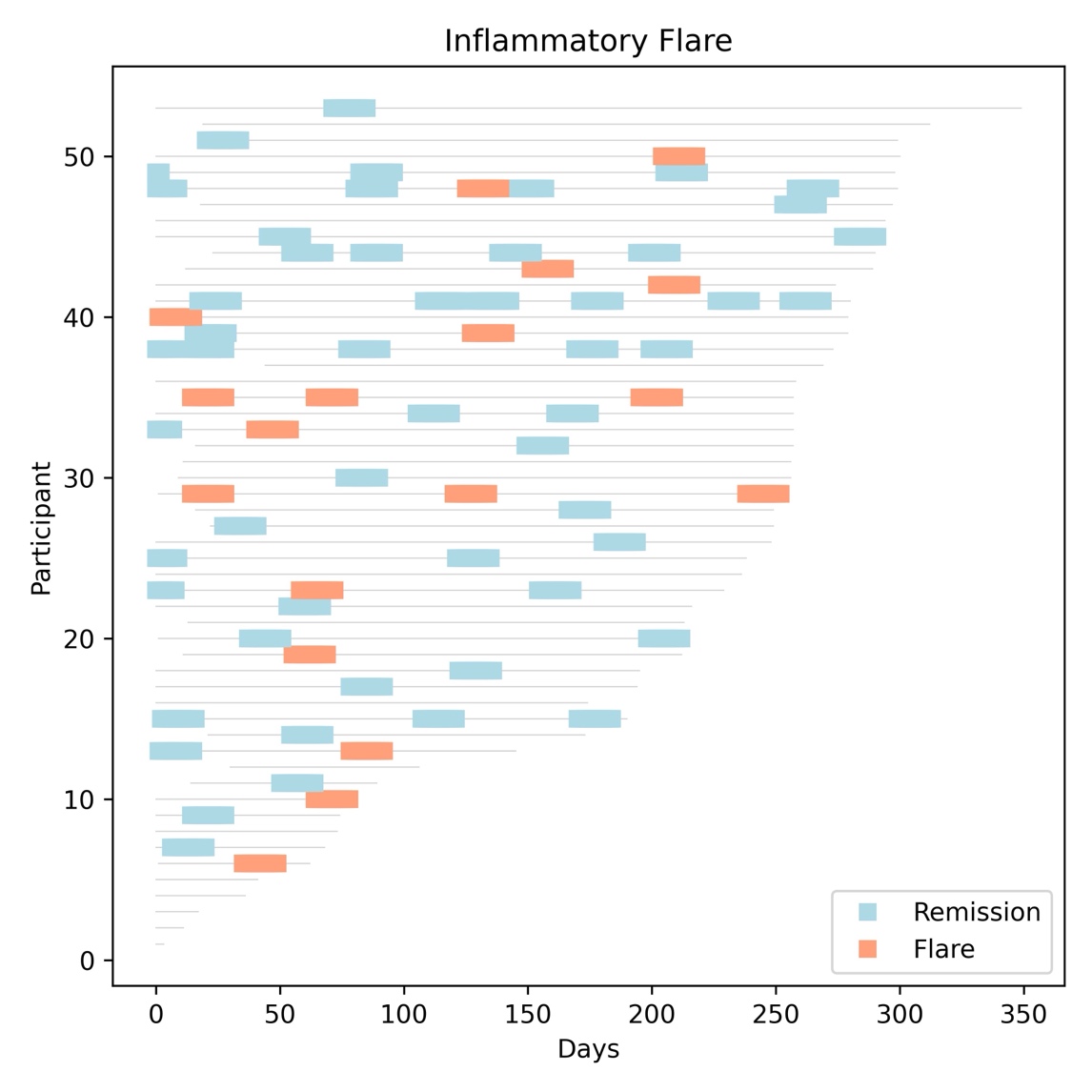

Supplemental Figure 1. The course of inflammatory activity for each participant over the study period. Inflammatory activity was assessed by C-reactive protein that was collected by each participant as part of their standard of care assessments. Values were imputed in a +/- 7-day period around each laboratory assessment. The y-axis indicates each study participant, ordered by number of days with any data (wearable/flare) contribution towards the study, and shown by the gray line. The x-axis indicates each day of the study that the participant was followed.

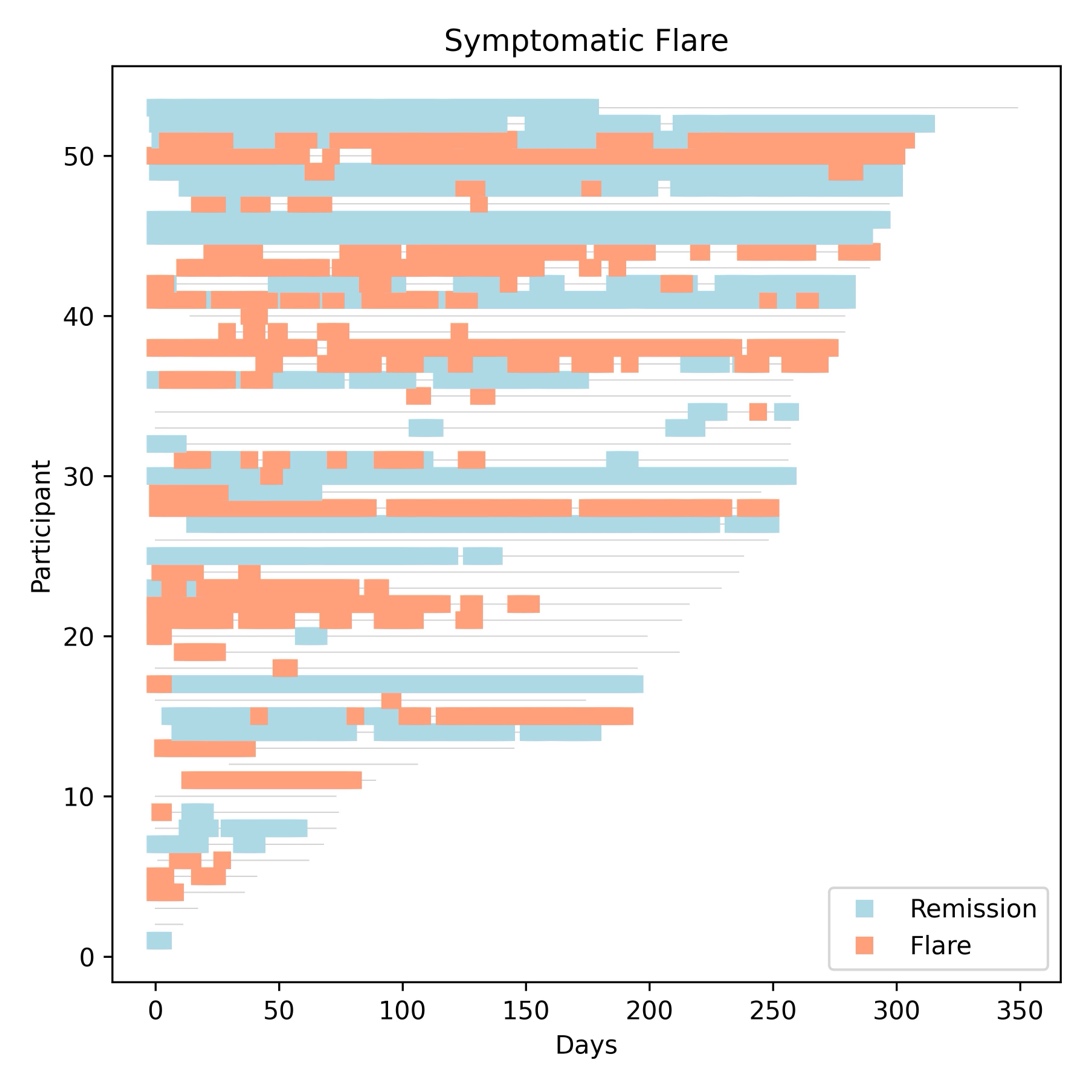

Supplemental Figure 2. The course of symptomatic activity for each participant over the study period. Symptomatic activity was assessed by the daily Routine Assessment of Patient Index Data (RAPID-3) scale. The y-axis indicates each study participant, ordered by number of days with any data (wearable/flare) contribution towards the study, and shown by the gray line. The x-axis indicates each day of the study that the participant was followed.

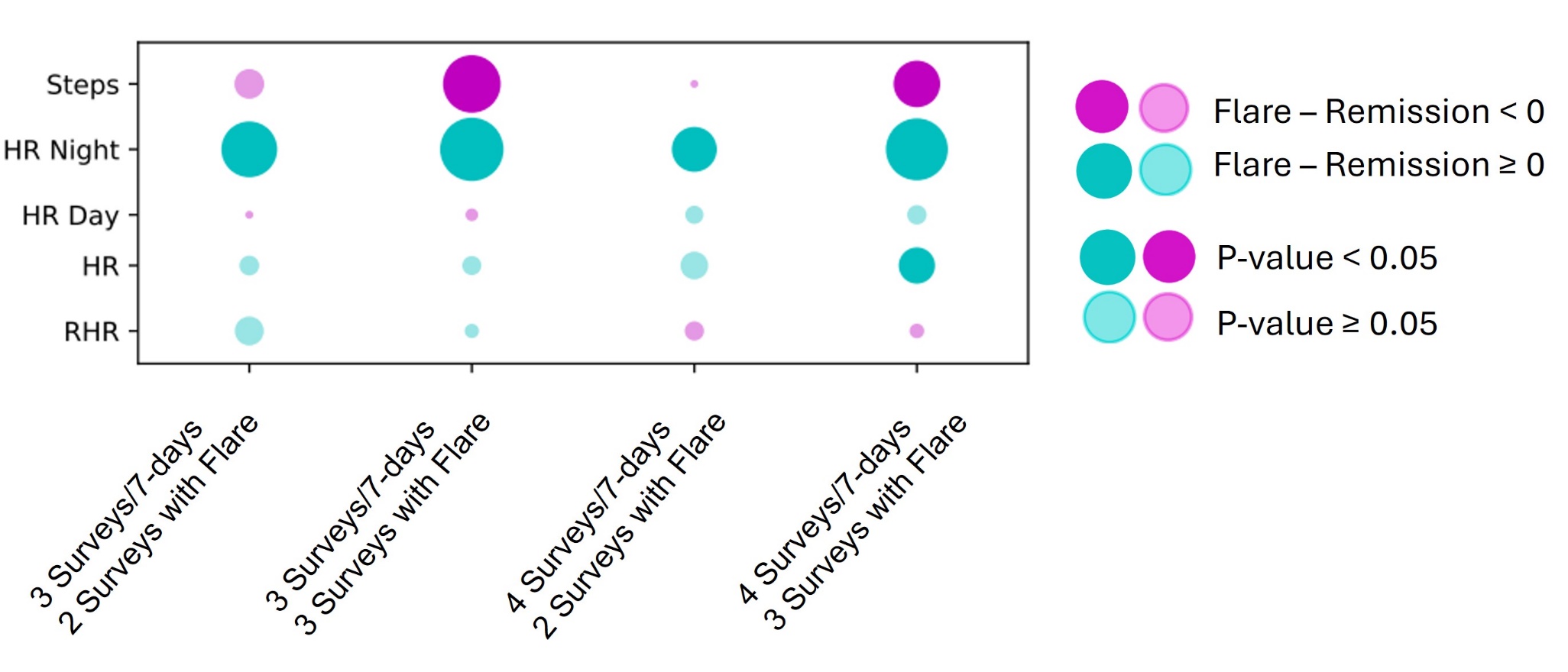

Supplemental Figure 3. The relationship between various symptomatic flare criteria and physiological metrics was explored. Steps, nighttime heart rate, daytime heart rate, daily heart rate and resting heart rate were calculated and associated with the daily Routine Assessment of Patient Index Data (RAPID-3) scale. Different combinations of the number of questionnaires answered per 7-day period and the number of daily survey scores meeting criteria for symptomatic flare in each 7-day period are shown. The strength and directionality of the statistical relationship between physiological metrics and survey results are denoted by the size and shading of each circle. Green indicates an increase in the metric during period of flare. Purple indicates a decrease in the metric during period of flare. Darker colors represent a statistically significant change.

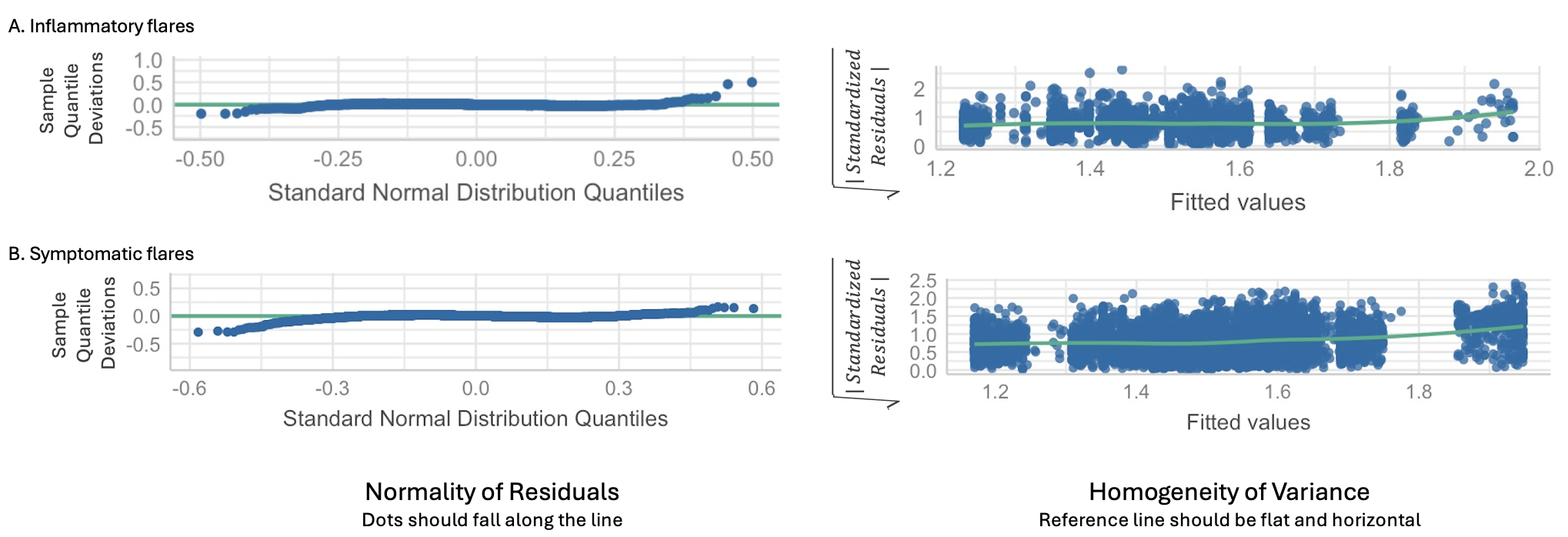

Supplemental Figure 4. HRV RMSSD cosinor model goodness-of-fit analysis for inflammatory (A) and symptomatic (B) flares showing residual plots with approximately normal distribution and homoscedasticity.

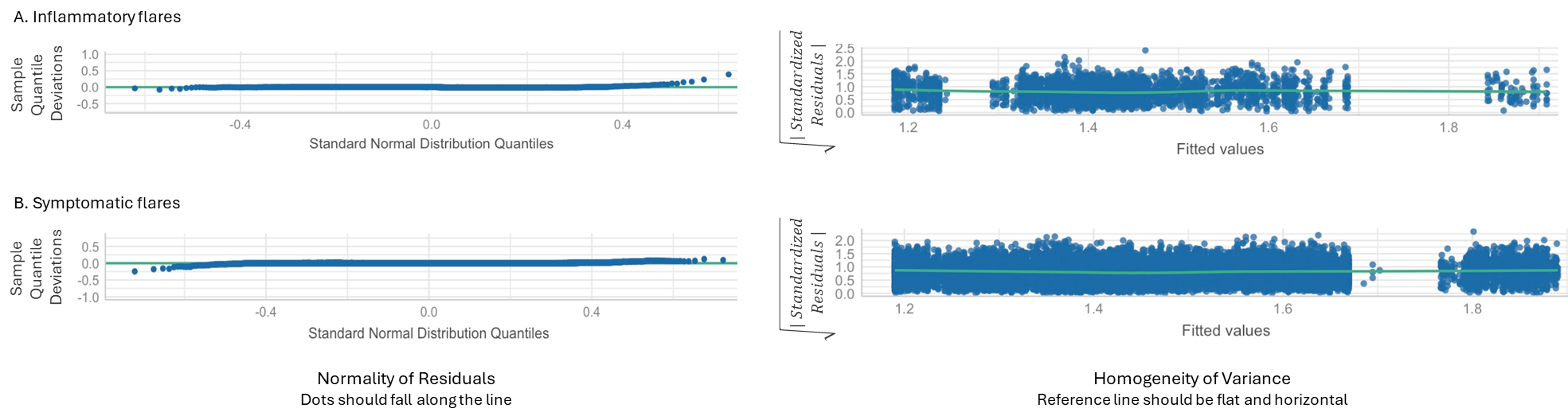

Supplemental Figure 5. HRV SDNN cosinor model goodness-of-fit analysis for inflammatory (A) and symptomatic flares (B) showing residual plots with approximately normal distribution and homoscedasticity.
